# Male gender and kidney illness associated with an increased risk of severe laboratory-confirmed coronavirus disease

**DOI:** 10.1101/2020.06.29.20142562

**Authors:** Efrén Murillo-Zamora, Xóchitl Trujillo, Miguel Huerta, Mónica Ríos-Silva, Oliver Mendoza-Cano

**Affiliations:** Departamento de Epidemiología, Unidad de Medicina Familiar No. 19, Instituto Mexicano del Seguro Social, Av. Javier Mina 301, Col. Centro, C.P. 28000, Colima, Colima, Mexico.; Centro Universitario de Investigaciones Biomédicas, Universidad de Colima, Av. 25 de julio 965, Col. Villas San Sebastián, C.P. 28045 Colima, México.; Universidad de Colima - Cátedras CONACyT, Centro Universitario de Investigaciones Biomédicas, Av. 25 de julio 965, Col. Villas San Sebastián, C.P. 28045 Colima, México.; Facultad de Ingeniería Civil, Universidad de Colima, km. 9 carretera Colima-Coquimatlán, Col. Jardines del Llano, C.P. 28400, Coquimatlán, Colima, Mexico.

**Keywords:** COVID-19, Severe acute respiratory syndrome coronavirus 2, Humans, Male, Kidney diseases

## Abstract

**Objective:** To identify factors predicting severe coronavirus disease 2019 (COVID-19) in adolescent and adult patients with laboratory-positive (quantitative reverse-transcription polymerase chain reaction) infection.

**Methods:** A retrospective cohort study took place, and data from 740 subjects, from all 32 states of Mexico, were analyzed. The association between the studied factors and severe (dyspnea requiring hospital admission) COVID-19 was evaluated through risk ratios (RRs) and 95% confidence intervals (CIs).

**Results:** Severe illness was documented in 28% of participants. In multiple analysis, male gender (RR = 1.13, 95% CI 1.06 - 1.20), advanced age ([reference: 15 - 29 years old] 30 - 44, RR = 1.02, 95% CI 0.94 - 1.11; 45 - 59, RR = 1.26, 95% CI 1.15 - 1.38; 60 years or older, RR = 1.44, 95% CI 1.29 - 1.60), chronic kidney disease (RR = 1.31, 95% CI 1.04 - 1.64) and thoracic pain (RR = 1.16, 95% CI 1.10 - 1.24) were associated with an increased risk of severe disease.

**Conclusions:** To the best of our knowledge, this is the first study evaluating predictors of COVID-19 severity in a large subset of the Latin-American population. It is also the first in documenting gender-related differences regarding the severity of the illness. These results may be useful for health care protocols for the early detection and management of COVID-19 patients that may benefit from opportune and specialized supportive medical treatment.

## Introduction

Since its first detection in December 2019,^1^ the coronavirus disease 2019 (COVID-19) caused by SARS-COV-2 (severe acute respiratory syndrome coronavirus 2) has spread rapidly to most regions of the world, including America. By April 9, 2020, nearly 1.5 million confirmed cases, and 85,522 deaths (6%, fatality rate) were registered globally.^2^

The COVID-19 pandemic represents a major challenge for healthcare systems worldwide, and published data evaluating factors associated with the severity of the disease are scarce.^3-5^ Identifying factors to predict disease severity in cases is fundamental to improve the survival rate from COVID-19.4 We aimed to identify what factors are associated with the risk of severe laboratory-confirmed COVID-19 among adolescent and adult patients in Mexico.

## Methods

### Study design

A retrospective cohort study was conducted in April 2020. Potential eligible subjects (laboratory-confirmed cases of COVID-19, quantitative reverse-transcription polymerase chain reaction, RT-qPCR) were identified from nominal records found in a national normative online system for respiratory viruses surveillance. Eligible cases were registered at any of more than 1,800 medical units (three levels of care) that the Mexican Institute of Social Security (*IMSS*, the Spanish acronym) has all across Mexico. The IMSS provides health care services to more than a third of the total population of Mexico.^6^

### Study population

Individuals aged 15 years or older, with symptoms onset from February 28 to March 27, 2020, and with conclusive test results (SARS-COV-2 infection confirmed) were eligible. Individuals with missing clinical or epidemiologic data of interest were excluded.

### Data collection

Demographic characteristics (sex, age), tobacco use (current), personal history of chronic communicable disease (HIV infection, no/yes) and noncommunicable disease (no/yes: obesity [body mass index of 30 or higher], arterial hypertension, type 2 diabetes mellitus, asthma, chronic kidney disease, immunosuppression, chronic obstructive pulmonary disease, or cardiovascular illness) were collected from the surveillance system. Date of healthcare-seeking and, when applicable, dates of hospital admission and discharge were also extracted from the audited database.

Additional clinical and epidemiologic data, such as if the influenza vaccine was applied during the same season as the onset of the acute illness (no/yes), and if acute symptoms were reported (cough, fever, headache, myalgia, arthralgia, odynophagia, chills, rhinorrhea, thoracic pain, diarrhea, polypnea, no/yes) were also extracted from the database. The primary data source were the medical records of the enrolled patients; these records were obtained from the employed surveillance resource system.

### Outcome

Laboratory-confirmed COVID-19 patients were classified as severe if they reported Dyspnea that resulted in hospital admission. Patients with severe manifestation were the main binary outcome (no/yes). Patients without dyspnea, despite being admitted to hospital, were considered as non-severe COVID-19 cases.

### Laboratory methods

According to normative standards,^7^ clinical specimens (nasopharyngeal or deep nasal swab) are analyzed (SuperScript™ III Platinum™ One-Step qRT-PCR Kits) at any of four specialized regional laboratories integrated with the *IMSS* network for epidemiologic surveillance. The laboratory methods employed in the *IMSS* network follow strict quality assurance standards in the diagnosis of viral respiratory pathogens.^8^

### Statistical analysis

Summary statistics were calculated, and the significance level was set at 5%. Risk ratios (RRs) and 95% confidence intervals (CIs), estimated by using generalized linear regression models, were employed to evaluate the association between the exposures to the analyzed risk, and the risk of severe COVID-19. All analyses were conducted using Stata version 14.0 (StataCorp).

### Ethical considerations

The Local Research Ethics Committee approved this study at the Mexican Institute of Social Security (IMSS, the Spanish acronym). With the Research Ethics Approval Number/ID: R-2020-601-015.

## Results

Data from 740 participants registered by 229 medical units located all across the country was analyzed. The study profile is shown in Figure 1. Dyspnea was registered in 285 individuals, and 207 of them required hospital admission. Therefore, they were classified as severe COVID-19 cases (28.0% from enrolled subjects).

**Figure 1.**
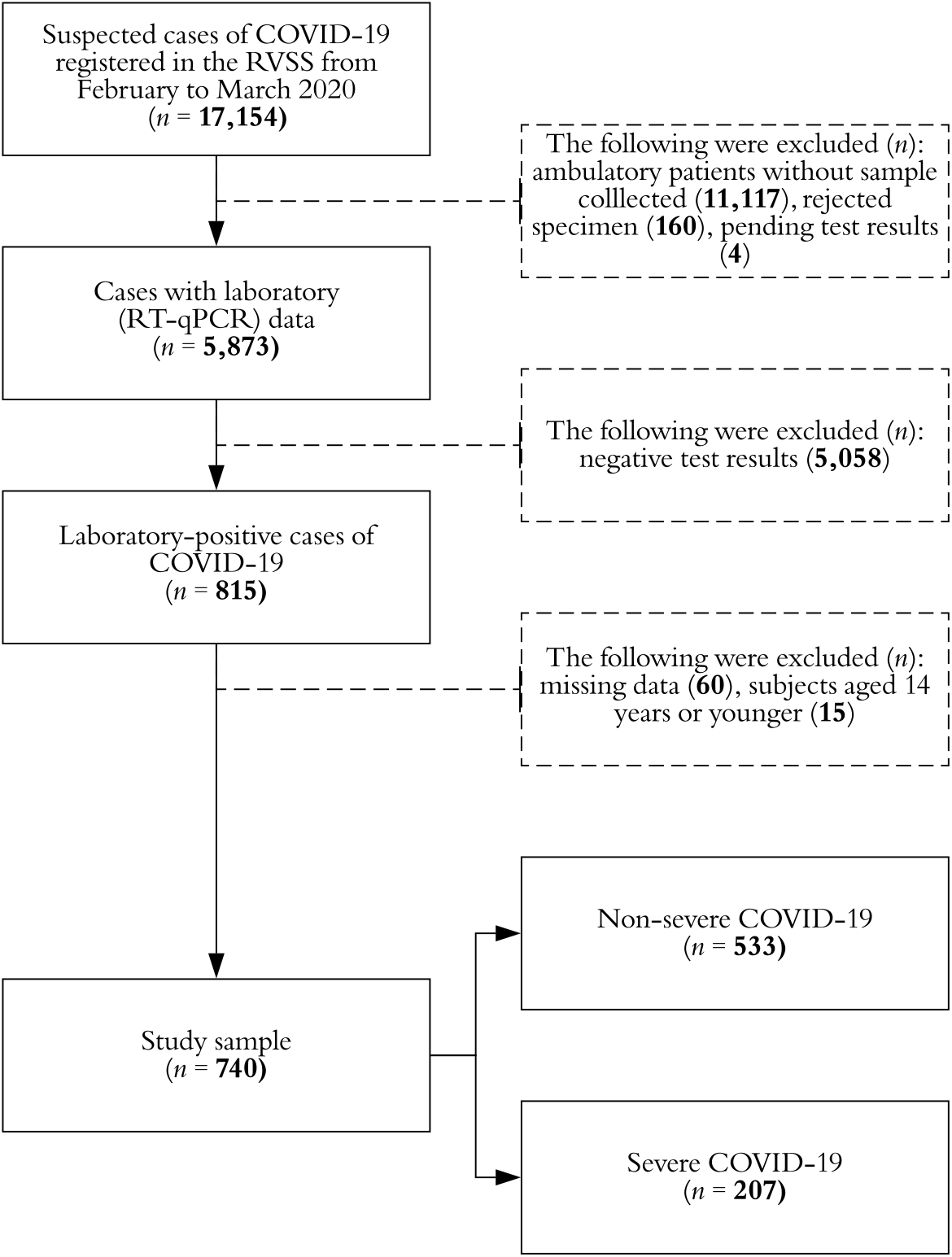
Study profile, Mexico 2020. Abbreviations: **RVSS**, Respiratory Virus Surveillance System; **COVID-19**, Coronavirus disease 2019; **RT-qPCR** (quantitative reverse-transcription polymerase chain reaction; SuperScript™ III Platinum™ One-Step qRT-PCR Kits in nasopharyngeal or deep nasal swabs).

Table 1 shows the characteristics of the study sample for the variables selected. Most of the participants were male (57.3%), and the overall mean age (± standard deviation) was 43.7 ± 14.9 years old. No gender-related differences were observed in terms of age (*p* = 0.371). Two-thirds of enrolled patients were aged 30 - 59 years old. Antipyretic drugs were prescribed to most (55.9%) subjects before they sought healthcare and, in 9 out of 10 them, acetaminophen was consumed. The mean length of hospital stay in severe cases was 4.9 ± 4.1 days and ranged from 0 to 19 days.

**Table 1.** Characteristics of analyzed individuals with laboratory-positive non-severe and severe COVID-19, Mexico 2020

|  | Overall<br>*n* = 740 |  | Non-severe<br>*n* = 533 |  | Severe<br>*n* = 207 |  | *p* |
| --- | --- | --- | --- | --- | --- | --- | --- |
| Sex |  |  |  |  |  |  |  |
| Female | 316 | (42.7) | 257 | (48.2) | 59 | (28.5) | < 0.001 |
| Male | 424 | (57.3) | 276 | (51.8) | 148 | (71.5) |  |
| Age (years) <sup>a</sup> | 43.7 ± 14.9 |  | 40.0 ± 13.0 |  | 53.1 ± 15.4 |  | < 0.001 |
| Age group (years) |  |  |  |  |  |  |  |
| 15 - 29 | 141 | (19.1) | 126 | (23.6) | 15 | (7.2) | < 0.001 |
| 30 - 44 | 281 | (38.0) | 240 | (45.1) | 41 | (19.8) |  |
| 45 - 59 | 210 | (28.3) | 122 | (22.9) | 88 | (42.6) |  |
| ≥ 60 | 108 | (14.6) | 45 | (8.4) | 63 | (30.4) |  |
| Received NAIs (any) |  |  |  |  |  |  |  |
| No | 734 | (99.2) | 529 | (99.2) | 205 | (99.0) | 0.769 |
| Yes | 6 | (0.8) | 4 | (0.8) | 2 | (1.0) |  |
| Received antyepitc drugs (any) |  |  |  |  |  |  |  |
| No | 326 | (44.1) | 227 | (42.6) | 99 | (47.8) | 0.198 |
| Yes | 414 | (55.9) | 306 | (57.4) | 108 | (52.2) |  |
| Antipyretic drug received <sup>b</sup> |  |  |  |  |  |  |  |
| Acetaminophen | 372 | (89.9) | 291 | (95.1) | 81 | (75.0) | < 0.001 |
| Metamizole | 11 | (2.7) | 5 | (1.6) | 6 | (5.6) |  |
| Ibuprofen | 10 | (2.3) | 4 | (1.3) | 6 | (5.6) |  |
| Other | 21 | (5.1) | 6 | (2.0) | 15 | (13.8) |  |
| Received influenza vaccine <sup>c</sup> |  |  |  |  |  |  |  |
| No | 622 | (84.1) | 440 | (82.6) | 182 | (87.9) | 0.073 |
| Yes | 118 | (15.9) | 93 | (17.4) | 25 | (12.1) |  |
| Elapsed days from symptoms onset to health-care seeking |  |  |  |  |  |  |  |
| < 1 | 347 | (46.9) | 253 | (47.5) | 94 | (45.4) | < 0.001 |
| 1 - 3 | 182 | (24.6) | 148 | (27.7) | 34 | (16.4) |  |
| 4 or above | 211 | (28.5) | 132 | (24.8) | 79 | (38.2) |  |
| Hospital admission |  |  |  |  |  |  |  |
| No | 470 | (63.5) | 470 | (88.2) | 0 | (0) | < 0.001 |
| Yes | 270 | (36.5) | 63 | (11.8) | 207 | (100) |  |
| **Acute symptoms profile** |  |  |  |  |  |  |  |
| Cough (yes) | 657 | (88.8) | 466 | (87.4) | 191 | (92.3) | 0.061 |
| Fever (yes) | 641 | (86.6) | 451 | (84.6) | 190 | (91.8) | 0.010 |
| Headache (yes) | 632 | (85.4) | 464 | (87.1) | 168 | (81.2) | 0.041 |
| Myalgia/Arthralgia (yes) | 572 | (77.3) | 423 | (79.4) | 149 | (72.0) | 0.031 |
| Odynophagia (yes) | 407 | (55.0) | 310 | (58.2) | 97 | (46.9) | 0.006 |
| Chills (yes) | 403 | (54.5) | 287 | (53.8) | 116 | (56.0) | 0.591 |
| Rhinorrhea (yes) | 353 | (47.7) | 276 | (51.8) | 77 | (37.2) | < 0.001 |
| Thoracic pain (yes) | 282 | (38.1) | 171 | (32.1) | 111 | (53.6) | < 0.001 |
| Diarrhea (yes) | 137 | (18.5) | 104 | (19.5) | 33 | (15.9) | 0.262 |
| Polypnea (yes) | 27 | (3.6) | 5 | (0.9) | 22 | (10.6) | < 0.001 |

Abbreviations: **COVID-19**, Coronavirus disease 2019; **NAIs**, Neuraminidase inhibitors; **COPD**, Chronic obstructive pulmonary disease; **HIV**, Human immunodeficiency virus.
Notes: 1) SuperScript™ III Platinum™ One-Step qRT-PCR Kits were used to confirm the COVID-19; 2) The absolute and relative (%) frequencies are presented, unless otherwise specified. 2) *p*-value from chi-square or t-tests as corresponding.
<sup>a</sup> The arithmetic mean ± standard deviation is presented.
<sup>b</sup> Among 414 subjects (non-severe disease, *n* = 306; severe disease, *n* = 108) in whom the use of antipyretic drugs was documented before healthcare seeking.
<sup>c</sup> During the same influenza season than acute COVID-19 onset.

**Table 1 (Cont.).** Characteristics of analyzed individuals with laboratory-positive non-severe and severe COVID-19, Mexico 2020
|  | Overall<br>*n* = 740 |  | Non-severe<br>*n* = 533 |  | Severe<br>*n* = 207 |  | *p* |
| --- | --- | --- | --- | --- | --- | --- | --- |
| ***Personal history of*** |  |  |  |  |  |  |  |
| Tobacco use (current, yes) | 80 | (10.8) | 47 | (8.9) | 32 | (15.6) | 0.009 |
| Obesity (BMI 30 or higher) (yes) | 162 | (21.9) | 100 | (18.8) | 61 | (29.7) | 0.001 |
| Arterial hypertension (yes) | 148 | (20.0) | 74 | (13.9) | 74 | (35.6) | < 0.001 |
| Cardiovascular disease (yes) | 20 | (2.7) | 8 | (1.5) | 12 | (5.9) | 0.001 |
| Type 2 diabetes mellitus (yes) | 108 | (14.6) | 51 | (9.5) | 58 | (27.8) | < 0.001 |
| Asthma (yes) | 33 | (4.4) | 22 | (4.2) | 10 | (4.9) | 0.676 |
| COPD (yes) | 27 | (3.7) | 9 | (1.7) | 18 | (8.8) | < 0.001 |
| Chronic kidney disease (yes) | 13 | (1.8) | 3 | (0.6) | 10 | (4.9) | < 0.001 |
| Immunosuppression (yes) | 19 | (2.5) | 8 | (1.5) | 10 | (4.9) | < 0.001 |
| HIV infection (yes) | 5 | (0.7) | 3 | (0.6) | 2 | (1.0) | 0.549 |

Abbreviations: **COVID-19**, Coronavirus disease 2019; **NAIs**, Neuraminidase inhibitors; **COPD**, Chronic obstructive pulmonary disease; **HIV**, Human immunodeficiency virus.
Notes: 1) SuperScript™ III Platinum™ One-Step qRT-PCR Kits were used to confirm the COVID-19; 2) The absolute and relative (%) frequencies are presented, unless otherwise specified. 2) *p*-value from chi-square or t-tests as corresponding.
<sup>a</sup> The arithmetic mean ± standard deviation is presented.
<sup>b</sup> Among 414 subjects (non-severe disease, *n* = 306; severe disease, *n* = 108) in whom the use of antipyretic drugs was documented before healthcare seeking.
<sup>c</sup> During the same influenza season than acute COVID-19 onset.

Severe COVID-19 cases, were older (53.1 ± 15.4 vs. 40.0 ± 13.0, *p* < 0.001) when compared with non-severe patients (Table 1), and particularly in the eldest age group (≥ 60 years old; 30.4% vs. 8.4%). Severe cases were also more likely to seek healthcare after four or more days after symptoms onset (38.2% vs. 24.8%, *p* < 0.001).

Regarding the acute symptoms profile, severe cases had a higher frequency of fever (91.8% vs. 84.6%, *p* = 0.010), thoracic pain (53.6% vs. 32.1%, *p* < 0.001) and polypnea (10.6% vs. 0.9%, *p* < 0.001). Significant and higher prevalences of pain-related symptoms (headache, myalgia, arthralgia, odynophagia) were documented in non-severe cases (Table 1).

In the multiple regression analysis (Table 2), male gender (RR = 1.13, 95% CI 1.06 - 1.20) and older patients ([reference: 15 - 29 years old] 30 - 44, RR = 1.02, 95% CI 0.94 - 1.11; 45 - 59, RR = 1.26, 95% CI 1.15 - 1.38; 60 years or older, RR = 1.44, 95% CI 1.29 - 1.60), subjects to thoracic pain (RR = 1.16, 95% CI 1.10 - 1.24) or chronic kidney disease (RR = 1.31, 95% CI 1.04 - 1.64) were also more likely to present severe COVID-19. On the other hand, in multiple analysis the history of obesity, tobacco use, cardiovascular or metabolic diseases, and pulmonary illness (namely asthma or chronic obstructive pulmonary disease) were not associated with the evaluated outcome.

**Table 2.**
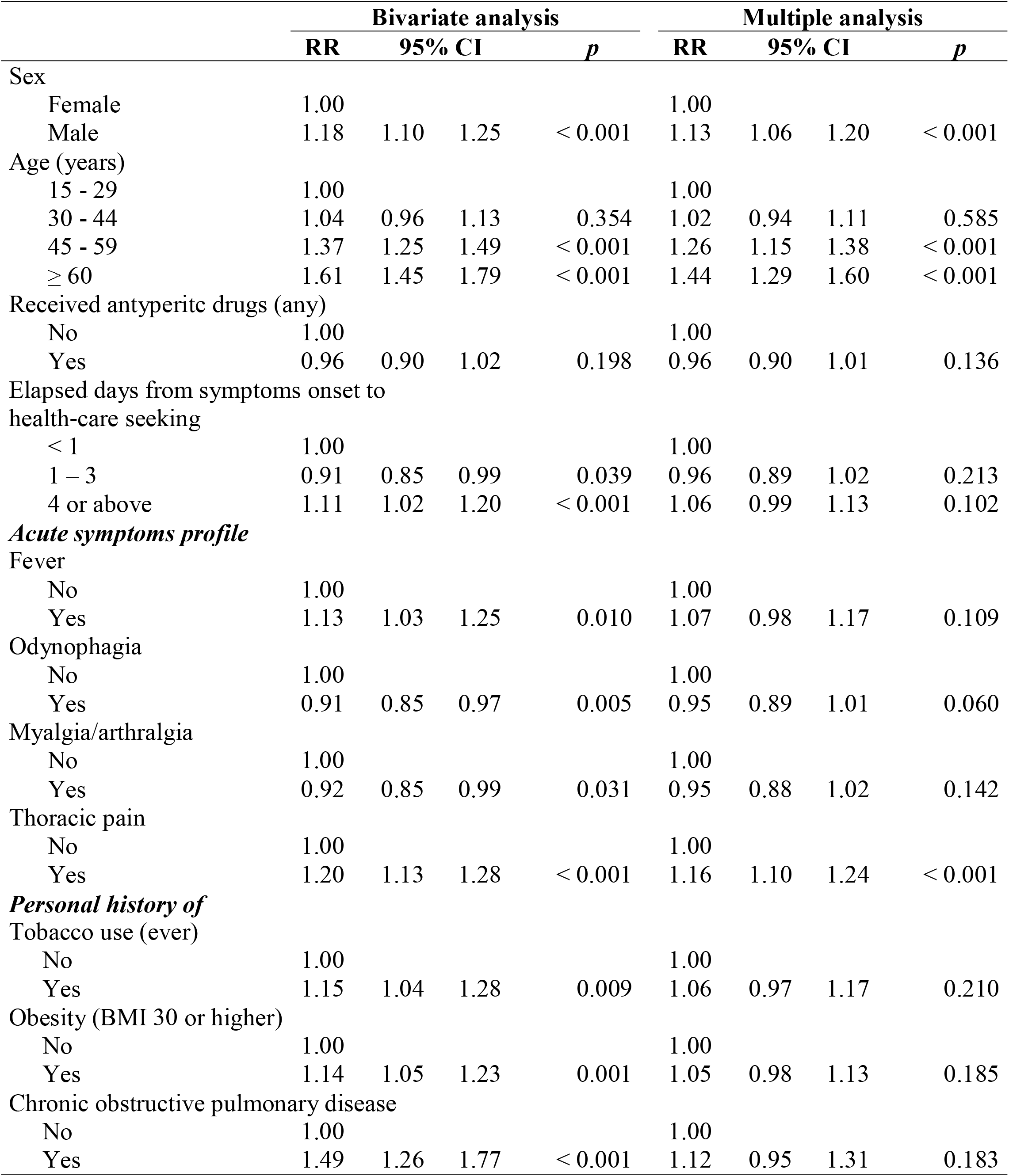
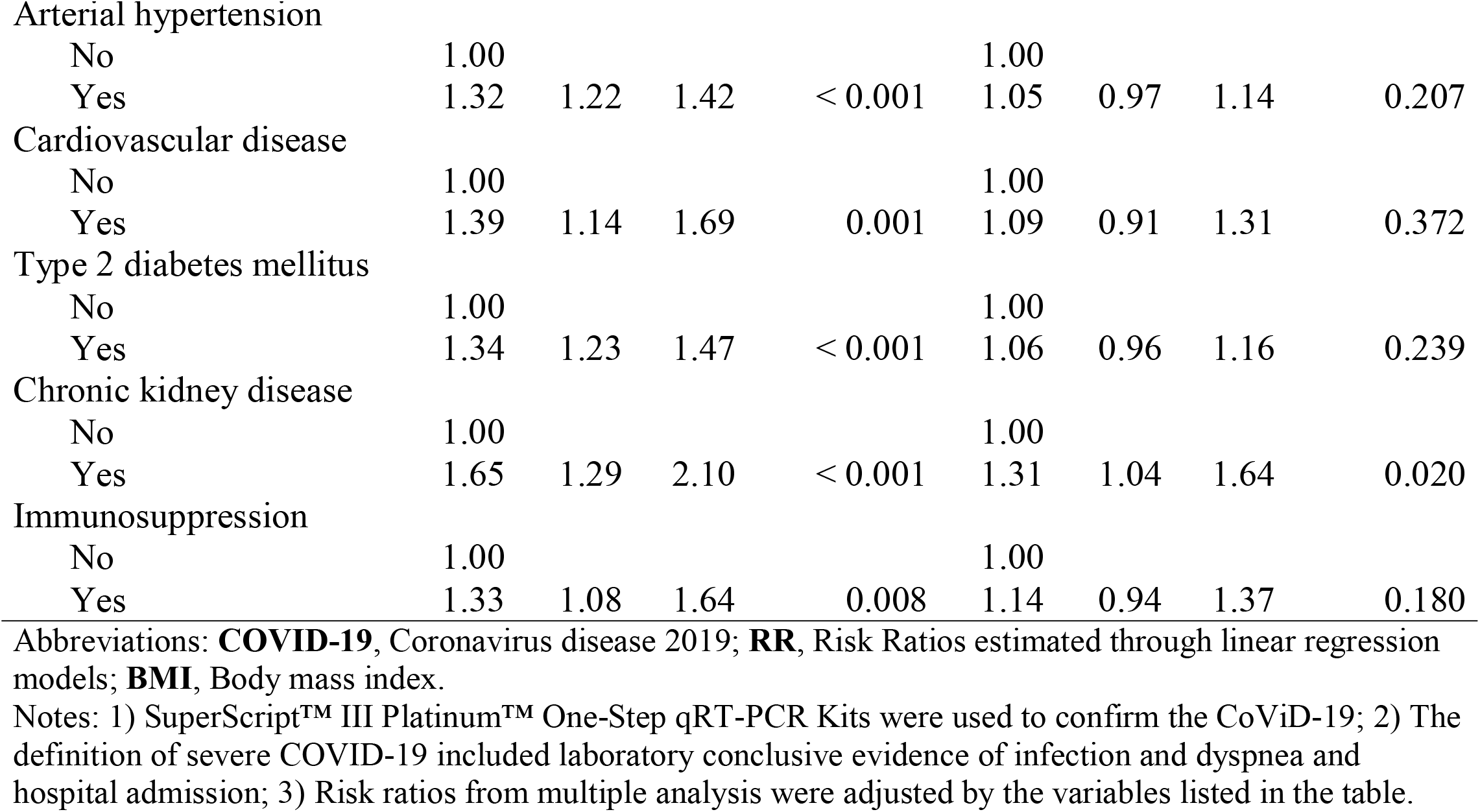
Factors associated with the risk of severe laboratory-confirmed COVID-19, Mexico 2020

## Discussion

Our findings suggest that more than 1 of 4 analyzed patients developed severe COVID-19 and its predictors were identified. To the best of our knowledge, this is the first study evaluating factors associated with the illness severity in a large subset of Latin-American subjects. This study is also the first, according to published scientific works, identifying gender-related differences in clinical outcomes of COVID-19 patients. These differences are biologically plausible.

Severe disease, according to our results, was presented by 28% (207/740) of enrolled laboratory-confirmed cases. This proportion is similar to government estimates (29%) at the national level (all healthcare institutions, including private facilities) by the end of the first week of April and when 3,181 RT-qPCR-confirmed cases had been registered.^9^

After adjusting by factors such as age, obesity and personal history of chronic noncommunicable diseases, males from our study sample were more likely (RR = 1.13, 95% CI 1.06 - 1.20) to develop dyspnea and to require hospital admission. Currently, the male gender is not considered a predictor for severe COVID-19.^5^ In a previously published analysis of 140 Chinese subjects with COVID-19, gender was not associated with an increased risk of severe illness.^3^ We hypothesize that sample size and differences in how a severe disease is defined, might be partially determining these discrepancies in the findings.

If later replicated, further research has to determine the pathogenic underlying mechanism between gender and the severity of SARS-COV-2 infection. Published data suggests that gender differences in susceptibility to SARS-COV (Severe Acute Respiratory Syndrome Coronavirus) in mice were secondary to estrogen receptor signaling, which seemed to be critical for protection in females.^10^ However, the role of sex hormones in the regulation of innate immune cells in the lung, as a response to viral respiratory pathogens, is poorly understood.^11^

In our study, a positive gradient between age and the risk of severe illness was observed (per additional year of age, RR = 1.010, 95% CI 1.007, 95% CI 1.012) in multiple regression analysis. Aging has been consistently associated with disease severity^3,12,13^ and its determining factors have not been described but may be related to viral load. A recently published analysis showed a positive and strong correlation (Spearman’s p = 0.48, 95% CI 0.074-0.75) between older age and RT-qPCR quantified viral load.^14^ Teenagers from our study sample (aged 15 – 19 years old, *n* = 5) seemed to have mild symptoms of SARS-COV-2 infection and all of them presented a non-severe form of disease.

Patients with previous medical diagnosis of chronic kidney disease had a 31% increase in the risk of severe COVID-19 (RR = 1.31, 95% CI 1.0 - 1.64). Interestingly, in our multiple regression study, neither type 2 diabetes mellitus (RR = 1.06, 95% CI 0.96 - 1.16) nor arterial hypertension (RR = 1.05, 95% CI 0.97-1.14), which are leading causes of renal impairment worldwide, were associated with the risk of severe COVID-19.

The association of previously diagnosed chronic kidney disease with the severity of COVID-19 was recently described in a subset of Chinese patients.^4^ When compared with national estimates (type 2 diabetes mellitus, 10.3%; arterial hypertension, 18.4%),^15^ higher prevalence of chronic noncommunicable diseases were observed in the study sample, particularly in participants with severe manifestations (type 2 diabetes mellitus, 27.8%; arterial hypertension, 35.6%).

Mexico lacks of a national registry of chronic kidney disease patients and its precise prevalence remains unknown.^16^ A previously published study where 3,564 randomly selected adults residing in urban areas of Mexico were analyzed, estimated that the population prevalence of chronic renal disease (creatinine clearance, Cockcroft-Gault formula) was 8% (< 60 ml/minute) and 0.1% (< 15 ml/minute) respectively.^17^

Thoracic pain was a common acute symptom and it was registered in medical records from 38.1% of enrolled subjects. It is a subjective symptom that seemed to be related to dyspnea, since it was presented by 55.1% and 27.5% of dyspneic and non-dyspneic patients respectively (*p* < 0.001).

The pathogenic mechanism of SARS-COV-2 in severe outcomes, including viral pneumonia, remains unclear; however, immunological changes seem to be crucial in the development of severe illness.^18^ Increased levels of proinflammatory cytokines, particularly interleukin 6, have been described among patients with severe COVID-19.^19^

Current tobacco use, obesity, and personal history of chronic obstructive pulmonary disease were only associated with the risk of severe disease in bivariate analysis. The overall prevalence of tobacco use (current) in the study sample was similar to the national mean (10.8% and 11.4%, respectively)^15^ and it was 37% higher among severe COVID-19 cases (15.6%). In China, there was a higher frequency for severe patients to be current smokers (16.9% vs. 11.8%), however, no association analysis was performed.^20^

Obesity is assessed by the analyzed surveillance system as a dichotomous variable (BMI equal or higher than 30, no/yes) and the current consensus is that morbidly obese induvials (BMI ≥ 40) are at increased risk of severe COVID-19.^5^ Since the patients’ height and usual weight are not collected by the surveillance system, we were unable to compute de BMI and therefore to estimate the prevalence of severe obesity in the study sample.

The potential limitations of our study must be cited. First, only users from a healthcare institution were enrolled (*IMSS*) and their characteristics may not be entirely representative from the source population. However, the profile of its users remains heterogeneous. Second, the system that served as source of data focus in epidemiological surveillance and clinical data (i.e. oxygen saturation) is not systematically collected. In addition, most of collected data is dichotomous (no/yes) in order to simplify its operation and we were unable to obtain other clinical and epidemiological data of interest.

## Conclusions

The COVID-19 pandemic is a major public health issue across the world. An effective response of healthcare systems is needed, which must include early identification of patients who are at increased risk of severe symptoms and poorer disease outcomes. To the best of our knowledge, this is the first study evaluating predictors of COVID-19 severity in a large subset of the Latin-American population. It is also the first documenting gender-related differences in the severity of illness. These results may also be useful in medical decision making related to the start of SARS-COV-2 antiviral therapy, when available.

## Data Availability

The data that support the findings of this study are available from the corresponding author, [MC, O], upon reasonable request.

## Ethics approval and consent to participate

Local IMSS Health Research Ethics Committee, with Approval code: R-2020-601-015.

## Consent for publication

Not applicable.

## Author Contributions

Efrén Murillo-Zamora conceived and designed the experiments also wrote the first draft of the manuscript; Xochitl Trujillo made data analysis and data collection, Miguel Huerta contributed with the Methodology and Writing—review and editing; Mónica Ríos-Silva contributed with revisions and data analysis and Oliver Mendoza-Cano performed the experiments, analyzed the data, and is responsible of the final version of the manuscript.

## Acknowledgments

The researchers would like to thank all the practicing physicians of family medicine that participated in data collections.

## Availability of data and materials

All data generated or analyzed during this study are included in this published article and its supplementary information files.

